# Spatial variability of nitrogen dioxide and formaldehyde and residential exposure of children in the industrial area of Viadana, Northern Italy

**DOI:** 10.1101/2020.04.16.20067777

**Authors:** Alessandro Marcon, Silvia Panunzi, Massimo Stafoggia, Chiara Badaloni, Kees de Hoogh, Linda Guarda, Francesca Locatelli, Caterina Silocchi, Paolo Ricci, Pierpaolo Marchetti

**Author notes:** **Corresponding author** Alessandro Marcon, PhD, Unit of Epidemiology and Medical Statistics, Department of Diagnostics and Public Health, University of Verona, c/o Istituti Biologici II, strada Le Grazie 8, 37134 Verona, Italy. **Contributorship statement** AM and PR are co-principal investigators of the Viadana III study. AM conceived the present analysis and drafted the first version of the manuscript. SP performed the geographical and statistical analysis. PM supervised the geographical and statistical analyses. CS and PR obtained data from the environmental protection agency (ARPA Lombardia). LG reconstructed the residential history of participants. CB and MS provided exposure estimates from EPISAT. KdH provided exposure estimates from ELAPSE. All the authors contributed in the discussion of the analysis plan, the interpretation of results, and commented on the first draft, they critically reviewed and approved the final version of the manuscript.

## Abstract

Chipboard production is a source of ambient air pollution. We assessed the spatial variability of outdoor pollutants and residential exposure of children living in proximity to the largest chipboard industry in Italy, and evaluated the reliability of exposure estimates obtained from a number of available models.

We obtained passive sampling data on NO_2_ and formaldehyde collected by the environmental protection agency of Lombardia region at 25 sites in the municipality of Viadana during 10 weeks (2017-18), and compared NO_2_ measurements with average weekly concentrations from continuous monitors. We compared interpolated NO_2_ and formaldehyde surfaces with previous maps for 2010. We assessed the relationship between residential proximity to the industry and pollutant exposures assigned using these maps, as well as other available countrywide/continental models based on routine data on NO_2_, PM_10_, and PM_2.5_.

The correlation between NO_2_ concentrations from continuous and passive sampling was high (Pearson’s r=0.89), although passive sampling underestimated NO_2_ especially during winter. For both 2010 and 2017-18, we observed higher NO_2_ and formaldehyde concentrations in the south of Viadana, with hot-spots in proximity to the industry. PM_10_ and PM_2.5_ exposures were higher for children at <1km compared to the children living at >3.5 km to the industry, whereas NO_2_ exposure was higher at 1-1.7 km to the industry. Road and population densities were also higher close to the industry.

Findings from a variety of exposure models suggest that children living in proximity to the chipboard industry in Viadana are more exposed to air pollution, and that exposure gradients are relatively stable over time.

## 1. INTRODUCTION

Industrial wood manufacturing and chipboard production are a source of ambient air pollutants, including dust from mechanical woodworking, formaldehyde from the resins used to bond wood particles, and combustion by-products (Dahlgren et al. 2003; Marcon et al. 2014).

The largest industrial park for the production of chipboard in Italy is located in the health district of Viadana (labelled “district” for short). The district is located in the Po Plain, in Northern Italy, one of the most polluted regions in Europe (Larsen et al. 2012; Stafoggia et al. 2017). It extends over an area of 363 km^2^ and comprises Viadana and 9 other smaller municipalities, counting 47,701 inhabitants overall in 2018 (http://demo.istat.it). The main emission sources of air pollution are two big industries in the south of the district, which include chemical plants for the synthesis of urea-formaldehyde resins, chipboard production and storage facilities, and small incinerators (Marcon et al. 2014). Both industries are under Directive 2010/75/UE for Integrated Pollution Prevention and Control.

We previously showed that living close to the industries is associated with several adverse outcomes in the paediatric population, including respiratory and irritation symptoms (de Marco et al. 2010; Girardi et al. 2012; Rava et al. 2012) and hospital admissions for respiratory diseases (Marchetti et al. 2014; Rava et al. 2011). Using data from *ad hoc* monitoring conducted by passive sampling in 2010, we found that residential proximity to the factories is linked to a higher outdoor exposure to nitrogen dioxide (NO_2_) and formaldehyde, and that exposure is associated with increased genotoxicity in mouth mucosa cells among children (Marcon et al. 2014).

NO_2_ has been related to a range of adverse effects, including mortality, exacerbations of obstructive airway diseases, and childhood asthma (Curtis et al. 2006). Formaldehyde is known to cause irritation and neuro-vegetative symptoms, and has mutagenic and carcinogenic properties (IARC 2006, update 2018). In urban environments, NO_2_ and formaldehyde mostly derive from combustions linked with road traffic, industrial activities, and domestic heating; formaldehyde is also generated through photo-oxidation of anthropogenic hydrocarbons (Kheirbek et al. 2012). In the district, wood waste incineration and power generation at the industrial premises are additional sources of nitrogen oxides, whereas formaldehyde is emitted during chipboard production and storage (Marcon et al. 2014).

The working principle of passive sampling is uptake of gaseous pollutants from the air by an adsorbing material at a constant uptake rate controlled by diffusion, followed by desorption and titration (Buczynska et al. 2009; Yu et al. 2008). As opposed to continuous monitors, passive samplers are cheap and easy to use since they do not require forced air movement through the device and thus a pump operated using electricity (Santana et al. 2017). However, diffusion rates can vary largely according to environmental conditions thus affecting their performance (Mason et al. 2011).

In the “Viadana III” study (http://biometria.univr.it/viadanastudy/) we are conducting new cohort studies of the population aged 0–21 years at baseline (2013), aimed to assess the relationship between exposure to air pollutants and further health outcomes during follow-up (2013–2017). The aim of the present analysis is to evaluate and compare possible exposure assignment methods to be used in the new follow-up study. To fulfil this aim:

1. we appraised the validity of passive sampling of NO_2_ against continuous monitoring (reference data were not available for formaldehyde);
2. we assessed whether higher air pollution concentrations in proximity to the industrial plants persisted over time, by comparing NO_2_ and formaldehyde surfaces obtained using recent (2017-18) monitoring data for the municipality of Viadana, where the largest chipboard industry is located, with our previous maps for 2010 (Marcon et al. 2014);
3. we evaluated the consistency between exposure estimates assigned through our maps and other available models that employed different input data, modelling techniques, and covered wider geographical areas. With the objective of focusing on residential areas, exposures were attributed to home addresses of Viadana III participants.

## 2. METHODS

### 2.1. Passive sampling monitoring campaigns

We obtained data from monitoring campaigns conducted using Radiello® passive samplers (Fondazione Salvatore Maugeri, Padova, Italy) both in 2010 and in 2017–2018.

The monitoring campaigns in 2010 were planned for epidemiological purposes and conducted in collaboration between our team and the environmental protection agency of Lombardia region (Agenzia Regionale per la Protezione dell’Ambiente [ARPA] Lombardia, Dipartimento di Cremona e Mantova, https://www.arpalombardia.it/). Briefly, measurements of formaldehyde and NO_2_ were conducted during two weeks in the cold season and two weeks in the warm season at 62 sites in the district (Marcon et al. 2014). Site selection overrepresented densely populated areas close to the two industries. Thirty-three of these sites were located in the eponymous municipality (labelled “Viadana” for short).

The monitoring campaigns conducted in 2017-18 were planned for institutional purposes by ARPA Lombardia. Our team did not participate in the selection of sites and periods. ARPA Lombardia identified twenty-five sites over a 10 km^2^ area around the centre of Viadana; none of these sites were the same as in 2010. NO_2_ and formaldehyde, but also BTEX (benzene, toluene, ethylbenzene, o-xylene, mp-xylene), acetaldehyde, and benzaldehyde were monitored during five weeks in the cold season (from 21/11/2017 to 27/12/2017) and five weeks in the warm season (from 8/5/2018 to 12/6/2018). When measurements were below the limit of quantification (LoQ), we assigned half the LoQ value, except when this occurred during all/most of the weeks (**Table S1**).

### 2.2. Data from continuous air quality stations

For 2017-18, we obtained hourly concentrations of NO_2_ from two air quality stations in Viadana (see maps in **Figure S1**) during the same periods of passive sampling. The first was a routine urban background station located 500 m away from the industry (via Gianmarco Cavalli, Viadana). The second was a temporary station located in a meadow close to a graveyard (via Cesare Airoldi, Viadana). One passive sampler was co-located with this station. The two stations were equipped with similar detectors in agreement with current legislation (D. Lgs. 155/2010), and measured NO_2_ by continuous chemiluminescence detectors. There were no traffic-type monitoring stations operating in the district area.

### 2.3. Geocoding of residential addresses

Viadana III includes prospective studies of two paediatric cohorts followed up between 2013 (baseline) and 2017. The first cohort is made of all the 3,854 children and adolescents born in 1992–2003 who were attending the schools in the district in 2006, when their parents were surveyed by questionnaire (de Marco et al. 2010). Of these, 3,711 (96%) subjects were traced through the healthcare provider records; 3,688 were still living in the district area in 2013. The second cohort included all the 4,221 children born in the district in 2004–2012 who lived in the district in 2013, and it was identified through electronic health records. Overall, the age range of the two cohorts was 0–21 years at baseline.

We geocoded residential addresses at baseline of participants in the two cohorts following the procedure described in **Appendix S1**. After excluding 206 and 178 addresses that failed to be geocoded from the first and second cohorts respectively, the number of available geocodes was 3,482 (94%) for the first cohort, and 4,043 (96%) for the second. We then combined coordinates from the two cohorts and removed duplicates (i.e. children living at the same address), obtaining a list of 4,390 unique locations in the district, 1,814 of which were in Viadana.

### 2.4. Population and street density indicators

With the aim of characterising non-industrial pollutant sources in the area, we obtained the number of inhabitants, buildings, and households per census tract by national statistics census data (ISTAT, 14° Censimento generale della popolazione e delle Abitazioni (http://dawinci.istat.it/), and derived population density indicators within 500 m buffers around the passive sampling locations by area-weighting. We also obtained the road network through Open Street Map (https://www.openstreetmap.org/), and derived total length of all types of roads in the 500 m buffers as an indicator of street density.

### 2.5. Exposure assignment

We obtained estimates of exposure to outdoor air pollutants at the 1,814 geocoded addresses by applying models available from three projects for baseline (or closest year), and new exposure models for 2017-18.

For baseline, we used the ordinary kriging models of NO_2_ and formaldehyde concentrations that we had previously devised using passive sampling data for 2010 (Marcon et al. 2014). For the same year, we also used the land-use regression (LUR) models for NO_2_, PM_2.5_, and black carbon (BC) concentrations developed for Western Europe within the ELAPSE study (Effects of Low-Level Air Pollution: A study in Europe) (de Hoogh et al. 2018). Dependent variables for these models were routine air quality data for NO_2_ and PM_2.5_ and *ad hoc* monitoring data for BC, whereas predictor variables included satellite data, dispersion model estimates, land cover and traffic indicators. Moreover, we obtained PM_10_ and PM_2.5_ concentrations for 2012 and 2013, respectively, using the spatiotemporal models developed for Italy within the EPISAT project (Dati satellitari ed uso del territorio per la stima delle esposizioni a livello nazionale) (Badaloni et al. 2018; Stafoggia et al. 2017). EPISAT models used routine air quality data as dependent variables, and incorporated both spatial (e.g. population density and emission data), and spatiotemporal (e.g. satellite data and meteorology) predictor variables. Co-authors MS and CB optimised exposure estimates for the study area aimed to capture PM variation due to very local sources, i.e. stage 4 modelling in Stafoggia et al. (2017).

For 2017-18, we devised new ordinary kriging models for NO_2_ and formaldehyde using passive sampling data. The best-fitting models were chosen by minimizing the root mean squared error (RMSE) by leave-one-out cross-validation (LOOCV) (Pebesma and Wesseling 1998). The spatial variogram providing the best model fit was a Gaussian class model for both pollutants. The variogram parameters were: partial sill = 13.4, range = 3.5 km, nugget = 2.5, and direction in plane = 135° (east) for NO_2_; and partial sill = 0.09, range = 2.0 km, nugget = 0.07 (no anisotropy) for formaldehyde. The estimated LOOCV-RMSEs were 1.95 and 0.28, respectively.

### 2.6. Statistical analysis

For the ten monitoring weeks in 2017-18, we calculated daily NO_2_ concentrations by averaging hourly data from each of the two continuous monitoring stations, setting to missing the days with less than 25% of hourly measurements available. Five (7%) daily concentrations were missing both for the routine and for the temporary station. Missing data can be non-randomly allocated over air pollution time-series, since they are typically generated by temporary breakdowns or instrumental maintenance. Therefore, we imputed missing data by k-nearest neighbour algorithm on the basis of available measurements from the two stations, as we did in Marchetti et al. (2017). We calculated weekly average concentrations using the imputed time-series. Then, by comparing weekly concentrations from the temporary station and the co-located passive sampler, we obtained a “recalibration equation” by linear regression. The resulting equation was used to predict NO_2_ concentrations at the 25 passive sampling sites, which are illustrated for the sake of comparison with national air quality standards.

We reported statistics on air pollution concentrations measured at the sampling sites, as well as exposures estimated at the residential addresses, as a function of distance to the industry. Sampling sites were categorised into three groups (<1.2, 1.2–1.7 and 1.8–15.5 km) whereas residential addresses were categorised into four groups (<1, 1–1.7, 1.8–3.4, 3.5–15.7 km) according to quantiles. Overall comparisons of exposures across groups were tested by the Kruskall-Wallis rank test.

The analyses were carried out using Stata Statistical Software, Release 16.1 (College Station, TX: StataCorp LLC) and R version 3.5.1 (http://www.R-project.org/).

### 2.7. Ethical approval

The Viadana III study has been approved by the Comitato Etico Val Padana (Prot. n. 4813, 12/02/2019).

## 3. RESULTS AND DISCUSSION

### 3.1. Comparison between passive sampling and continuous monitoring

All the NO_2_ measurements obtained using Radiello® tubes during the cold season, and most of those obtained in the warm season, were below the average concentrations derived by the continuous stations for the corresponding weeks (**Figure 1**). A comparison of weekly concentrations between the temporary station and the co-located sampler (solid and dashed thick lines, respectively) suggests that passive sampling underestimated NO_2_ concentrations especially in winter. In a validation study of different diffusive sampler types, Gerboles et al. (2006) found that Radiello® tubes provided lower NO_2_ concentrations compared to other samplers; Radiello® tubes underestimated reference concentrations both in a rural setting and in laboratory conditions, in particular when temperature, humidity and wind speed were set low. We observed a considerable variability across monitoring weeks, ranging from an overestimation on week 6 to a heavy underestimation on week 1, which supports the common approach of averaging concentrations across a number of monitoring periods before building exposure models, in order to level off measurement error (Beelen et al. 2013; de Hoogh et al. 2018). The correlation between the temporary station and the co-located sampler was high, as indicated by a Pearson’s r coefficient of 0.89 (**Figure S2**).This high correlation suggests that NO_2_ exposure estimates based on passing sampling data can be reasonably used to detect exposure-response relationships in the Viadana III epidemiological study. A caveat for the interpretation is that associations estimated using underestimated exposure metrics will be overestimated.

**Fig. 1.**
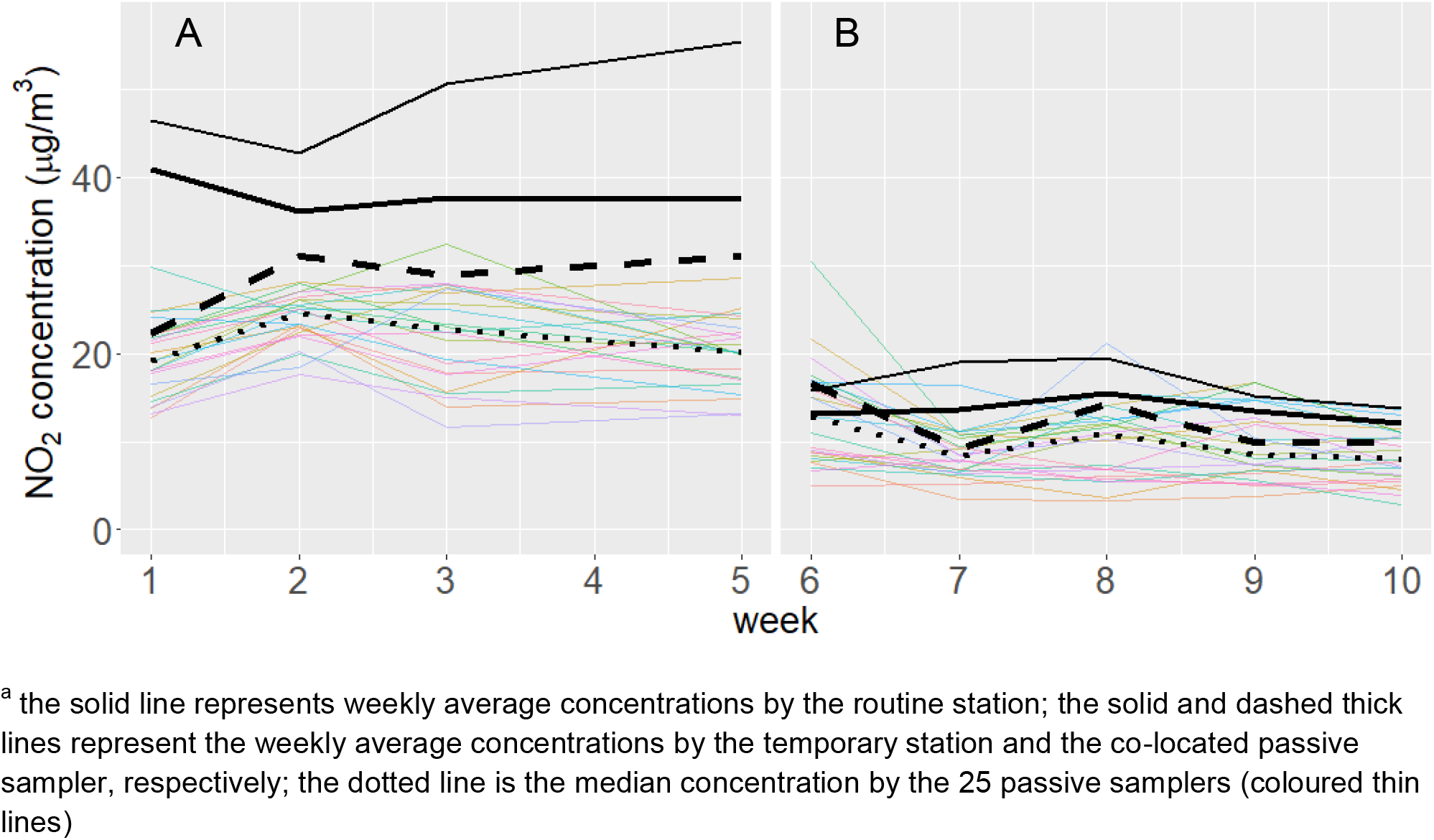
Weekly concentrations of NO_2_ measured by continuous monitors and passive samplers during the cold (panel A) and warm seasons (panel B). Municipality of Viadana, 2017-18.

For formaldehyde, since we did not have reference data and validation studies of passive sampling are few, comparison of concentrations with other studies should be done cautiously. Using Radiello® tubes, Evans and Stuart (2011) observed quite similar winter concentrations of formaldehyde between active monitors and co-located passive samplers (2.1 vs 2.2 µg/m^3^) in Hillsborough County, Florida. However, Mason et al. (2011) documented an excellent performance in controlled experiments (2% difference from a reference method) but substantial (39%) underestimation of formaldehyde in a real-world setting.

### 3.2. NO_2_ concentrations

The mean concentrations of NO_2_ during the cold and warm seasons in 2017-18 were 19.8 and 9.9 µg/m^3^, respectively (**Table 1**). Spatial variability was higher for NO_2_ during the warm season (coefficient of variation = 31%), compared to measurements during the cold season as well as to the other pollutants (**Table 2**). The median annual measured concentration was 16.0 (Q1–Q3: 12.4–16.6) µg/m^3^. After recalibration, annual concentrations were within the national air quality standard of 40 µg/m^3^ at all the sites (**Figure 2**), and the median annual concentration was 26.9 (Q1–Q3: 18.5–28.2) µg/m^3^. For comparison, average NO_2_ concentrations measured by routine stations in 2017-18 were around 30 µg/m^3^ for the urban area of Mantova, and 20 µg/m^3^ for the larger province encompassing also suburban/rural areas (ARPA Lombardia 2018).

**Table 1.** Distribution of weekly measurements of air pollutants from 25 passive samplers in Viadana, and daily temperature and rain precipitations from the nearest meteorological station for the same weeks (2017-18).

| Week n. | Starting date<br>(DD/MM/YY) | NO<sub>2</sub>, µg/m<sup>3</sup><br>(mean, SD) | Formaldehyde,<br>µg/m<sup>3</sup> (mean, SD) | Temperature, °C<br>(mean, min–max) | Rain, mm<br>(mean, min–max) |
| --- | --- | --- | --- | --- | --- |
| 1 | 21/11/17 | 19.5 (4.3) | 1.6 (1.0) | 6.8 (2.9–9.5) | 2.6 (0.0–11.4) |
| 2 | 28/11/17 | 24.1 (3.1) | 3.2 (0.6) | 2.8 (0.9–5.0) | 0.0 (0.0–0.0) |
| 3 | 05/12/18 | 22.3 (5.5) | 2.0 (1.5) | 1.7 (-0.8–4.3) | 1.7 (0.0–9.4) |
| 4 | 12/12/18 | 12.2 (5.6) | 2.6 (1.2) | 2.2 (-0.9–3.4) | 0.1 (0.0–0.4) |
| 5 | 18/12/18 | 20.8 (4.5) | 1.8 (0.9) | 1.3 (-0.6–5.2) | 0.0 (0.0–0.0) |
| Overall (cold season) |  | 19.8 (3.5) | 2.2 (0.5) | 2.9 (-0.9–9.5) | 0.8 (0.0–11.4) |
| 6 | 08/05/18 | 13.2 (5.9) | 0.6 (0.2) | 18.7 (16.1–20.4) | 2.2 (0.0–7.4) |
| 7 | 15/05/18 | 8.5 (2.7) | 0.7 (0.4) | 17.5 (12.8–19.7) | 0.7 (0.0–5.2) |
| 8 | 22/05/18 | 10.1 (4.3) | 0.6 (0.4) | 21.8 (17.3–23.9) | 7.8 (0.0–50.6) |
| 9 | 29/05/18 | 9.5 (3.9) | 0.8 (0.5) | 23.2 (22.4–23.8) | 0.4 (0.0–2.0) |
| 10 | 05/06/18 | 8.3 (2.9) | 2.6 (0.8) | 23.6 (20.5–25.1) | 5.0 (0.0–32.2) |
| Overall (warm season) |  | 9.9 (3.1) | 1.1 (0.3) | 21.0 (12.8–25.1) | 3.2 (0.0–50.6) |

**Table 2.** Distribution of measured concentrations of air pollutants at the 25 passive sampling locations in Viadana (2017-18), by groups based on tertiles of distance to the industry.

|  | Distance to the industrial plant |  |  |  | Coefficient of variation (%)<sup>b</sup> (n=25) |
| --- | --- | --- | --- | --- | --- |
|  | <1.2 km<sup>a</sup> (n=8) | 1.2–1.7 km<sup>a</sup> (n=9) | 1.8–15.5 km<sup>a</sup> (n=8) | Overall<sup>a</sup> (n=25) |  |
| NO<sub>2</sub> (cold season), µg/m<sup>3</sup> | 21.5 (19.8–23.6) | 19.4 (18.8–21.2) | 16.4 (14.7–20.4) | 19.9 (17.5–22.0) | 18 |
| NO<sub>2</sub> (warm season), µg/m<sup>3</sup> | 11.6 (9.5–13.4) | 9.0 (6.6–13.0) | 8.8 (6.5–11.0) | 9.9 (6.7–13.0) | 31 |
| NO<sub>2</sub> (annual), µg/m<sup>3</sup> | 16.4 (15.8–16.6) | 14.4 (13.0–17.1) | 11.9 (10.9–16.1) | 16.0 (12.4–16.6) | 19 |
| Formaldehyde (cold season), µg/m<sup>3</sup> | 2.3 (2.1–2.4) | 2.0 (1.71–2.7) | 2.2 (2.0–2.3) | 2.2 (2.0–2.6) | 22 |
| Formaldehyde (warm season), µg/m<sup>3</sup> | 1.1 (1.1–1.4) | 1.1 (0.9–1.2) | 1.0 (0.9–1.0) | 1.0 (0.9–1.2) | 25 |
| Formaldehyde (annual), µg/m<sup>3</sup> | 1.7 (1.5–2.0) | 1.5 (1.4–2.0) | 1.6 (1.4–1.7) | 1.6 (1.5–2.0) | 18 |
| Acetaldehyde (cold season), µg/m<sup>3</sup> | 0.7 (0.6–0.7) | 0.7 (0.6–0.9) | 0.7 (0.6–0.8) | 0.7 (0.6–0.8) | 19 |
| Benzene (cold season), µg/m<sup>3</sup> | 1.6 (1.5–1.6) | 1.6 (1.3–1.6) | 1.6 (1.5–1.6) | 1.6 (1.5–1.6) | 10 |
| Toluene (cold season), µg/m<sup>3</sup> | 2.9 (2.8–3.0) | 2.9 (2.5–3.1) | 2.8 (2.6–3.2) | 2.8 (2.6–3.1) | 11 |

<sup>a</sup> Medians with interquartile ranges reported.
<sup>b</sup> 100 × standard deviation/mean (%)

**Fig. 2.**
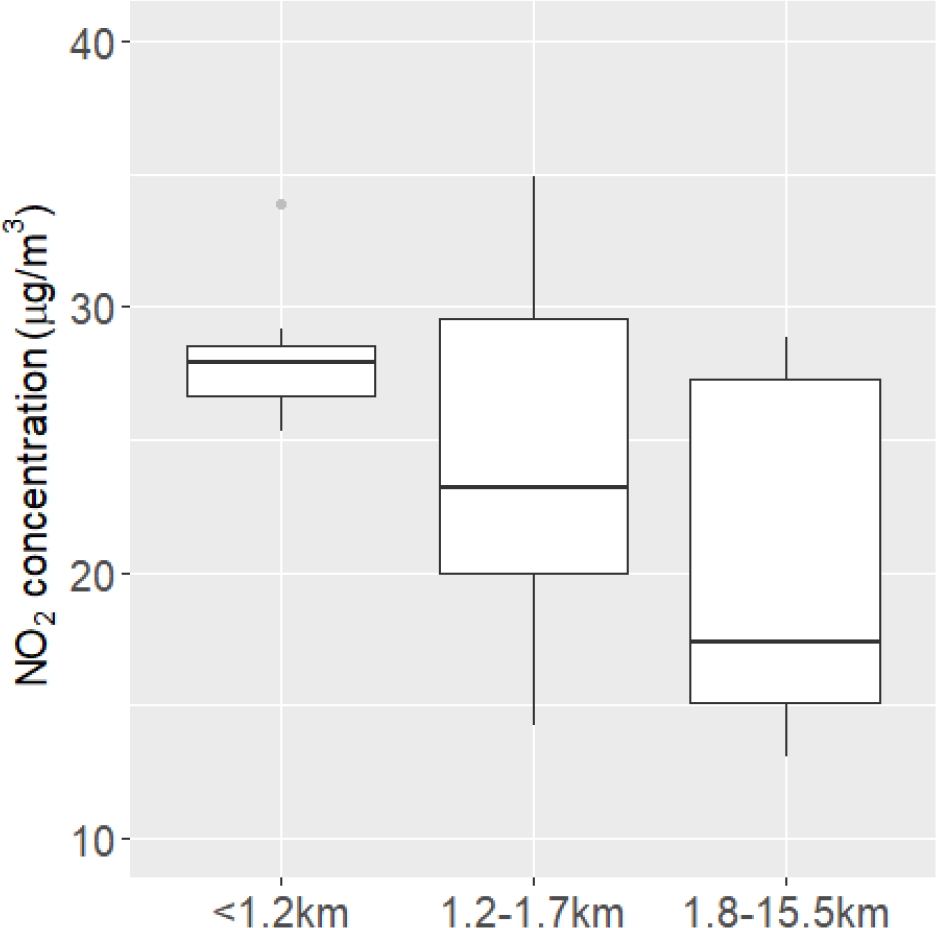
Box & whiskers plot of recalibrated NO_2_ concentrations at passive sampling locations in Viadana (2017-18), by groups based on tertiles of distance to the industry.

A clear relationship with distance was found when looking at observed NO_2_ concentrations (**Table 2**). The median recalibrated concentration was 11 µg/m^3^ higher at <1.2 km to the industry compared to the area >1.8 km (**Figure 2**).

### 3.3. Formaldehyde concentrations

As opposed to the campaigns in 2010 (Marcon et al. 2014), we found higher formaldehyde concentrations in the cold season than in the warm season (2.2 vs 1.1 µg/m^3^, respectively, **Table 1**), which was unexpected because lower concentrations are usually found in winter (Kheirbek et al. 2012; Villanueva et al. 2014). The median annual concentration of formaldehyde was 1.6 µg/m^3^ (Q1–Q3: 1.5–2.0 µg/m^3^), which is similar to other studies that used Radiello® tubes (Evans and Stuart 2011; Kheirbek et al. 2012). There are no ambient air quality standards for aldehydes, and studies on the health effects of outdoor formaldehyde are still few (Dahlgren et al. 2003; Marcon et al. 2014; Morello-Frosch et al. 2000). For long-term exposure, Effects Screening Levels (calculated so that lower ambient levels are unlikely to be of concern for health and the environment) are set to 3.3 µg/m^3^ from the Texas Air Monitoring Information System (USA – Texas Commission on Environmental Quality, https://www.tceq.texas.gov/ accessed 11 March 2020) (Santana et al. 2017). However, formaldehyde concentrations in our study were above 0.8 µg/m^3^, the benchmark set by the US Environmental Protection Agency for a 1 in 100,000 lifetime cancer risk for inhaled exposures (https://cfpub.epa.gov/ accessed 12 March 2020).

Observed concentrations did not vary appreciably according to proximity to the industry (**Table 2**), and spatial contrasts were small, as we observed during the previous campaigns (Marcon et al. 2014). Using Radiello® tubes, Kheirbek et al. (2012) reported a coefficient of variation of 22% for formaldehyde in New York City at springtime. Even lower coefficients of 12-13% were reported in other studies (Evans and Stuart 2011; Marcon et al. 2014).

### 3.4. Concentrations of other pollutants

In the cold season, mean concentrations of benzene, toluene, and acetaldehyde were 1.5, 2.8, and 0.7 µg/m^3^ respectively (**Table S3**), whereas they were mostly below the LoQ during the warm season (**Table S1**). Benzene and toluene concentrations were comparable or lower to concentrations observed at background or semirural sites in other studies using Radiello® tubes (Buczynska et al. 2009; Gaeta et al. 2016; Kerchich and Kerbachi 2012). Ethylbenzene, o-xylene, mp-xylene, and benzaldehyde concentrations were below the LoQ during both seasons.

### 3.5. Interpolated surfaces of NO_2_ and formaldehyde

For both NO_2_ (**Figure 3**) and formaldehyde (**Figure 4**), we observed higher concentrations in the south of Viadana, with hot-spots of concentrations above the 90^th^ percentile in proximity to the industry. However, higher concentrations were estimated for the north-eastern area of the municipality in 2010 compared to 2017-18 for both pollutants, which could have been driven by the different location of samplers between the two campaigns. Very few samplers were placed in that area, possibly affecting estimation accuracy and precision. The same holds true for the area at the East of the industry, where no samplers were located in 2017-18.

**Fig. 3.**
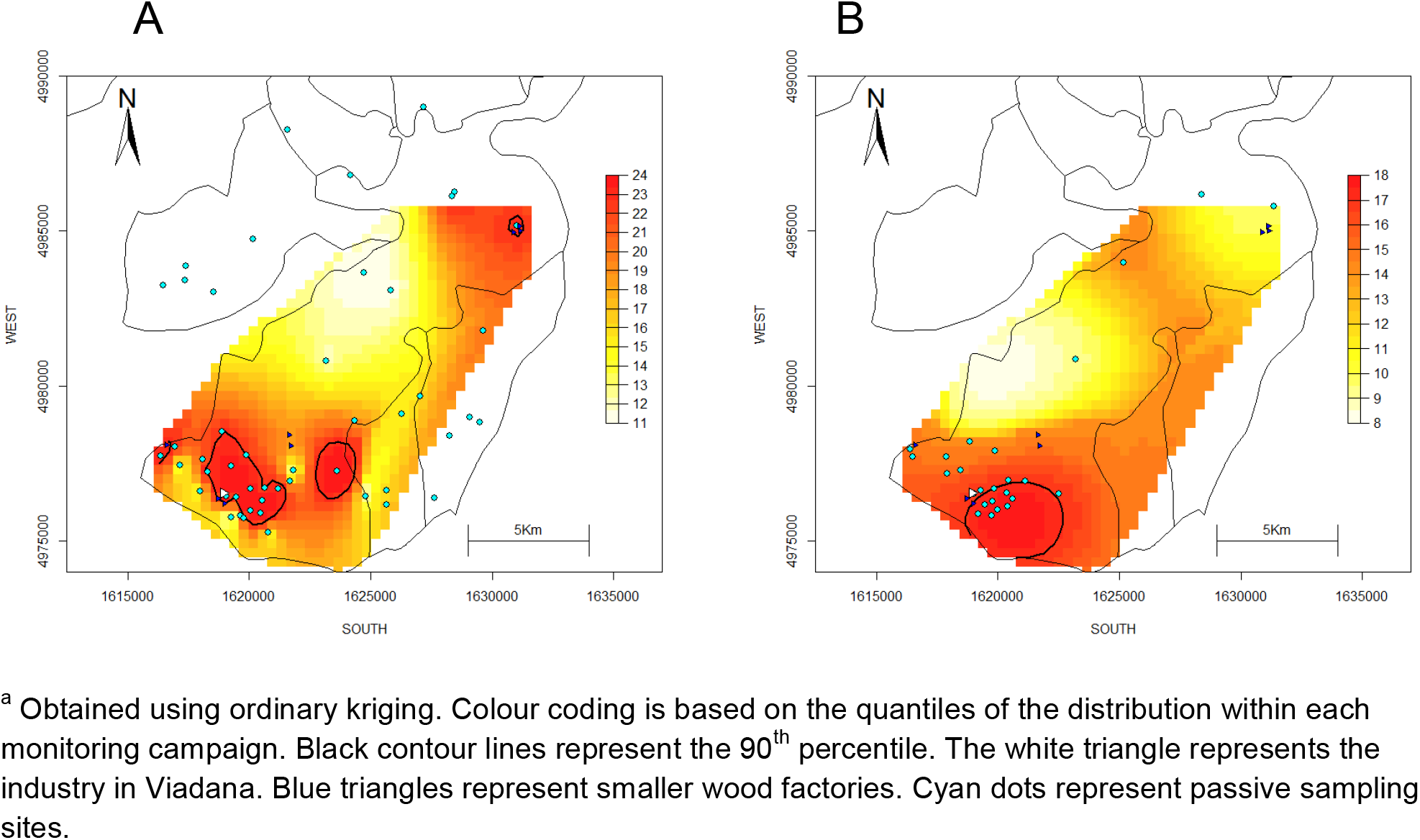
Spatial distribution of annual NO_2_ concentrations measured by passive samplers in 2010 (panel A) and 2017-18 (panel B).

**Fig. 4.**
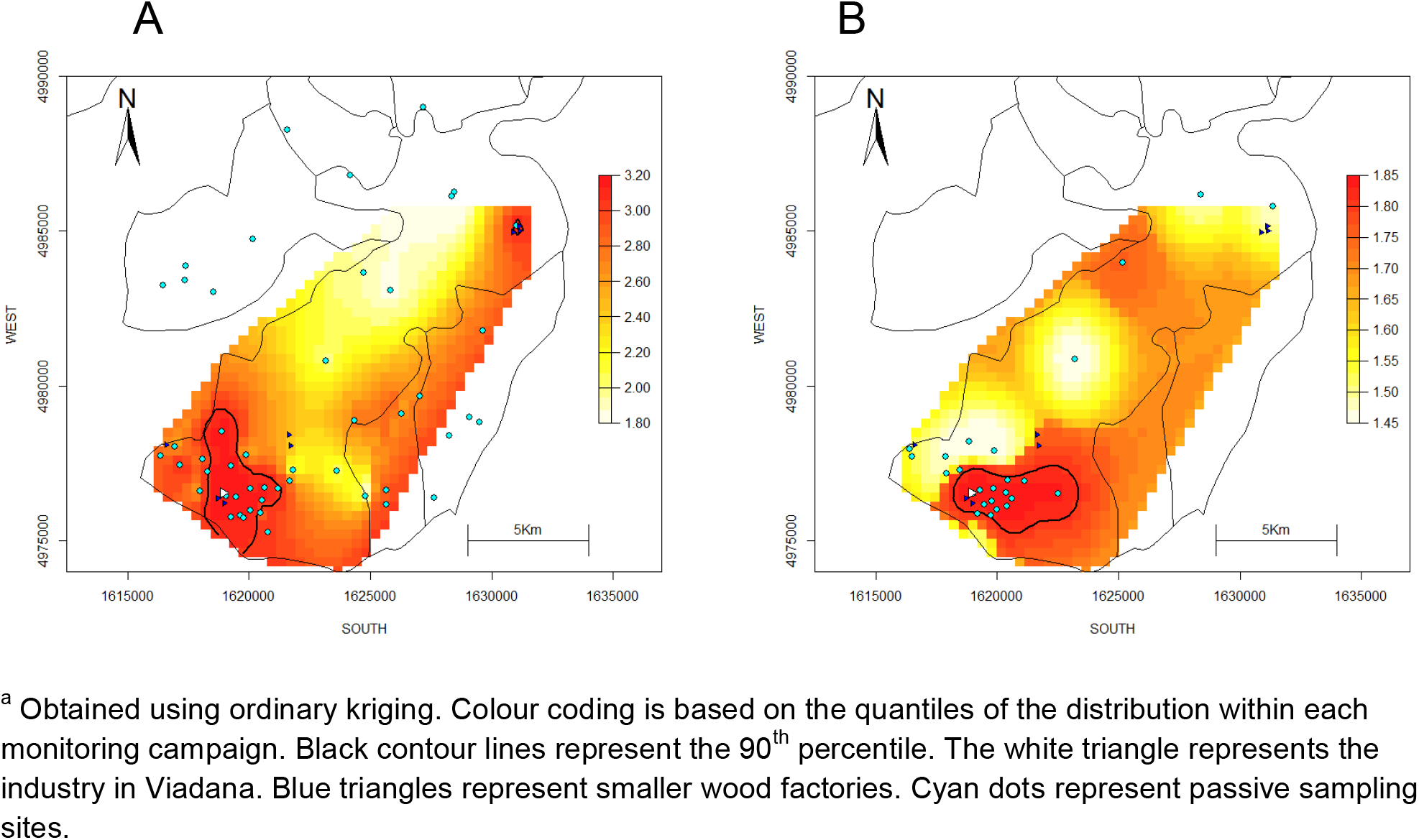
Spatial distribution of annual formaldehyde concentrations measured by passive samplers in 2010 (panel A) and 2017-18 (panel B).

Areas closer to the industry were characterised by a higher density of roads, population and buildings (**Table 3**). For example, the median total road length in 500-m buffers around the sampling sites decreased from 8.3 km to 4.6 km as the distance to the factory increased (from <1.2 to >1.8 km). It is therefore likely that road traffic and domestic heating contribute the higher levels of air pollution observed in proximity to the industry.

**Table 3.** Distribution of population and road density indicators in 500 m buffers around the 25 passive sampling sites in Viadana, by groups based on tertiles of distance to the industry.

|  | Distance to the industrial plant |  |  |  |
| --- | --- | --- | --- | --- |
|  | <1.2 km<br>(n=8) | 1.2–1.7 km<br>(n=9) | 1.8–15.5 km<br>(n=8) | Overall<br>(n=25) |
| Number of inhabitants | 1506 (1048–2701) | 713 (536–2702) | 527 (337–939) | 993 (536–2383) |
| Number of buildings | 332 (236–508) | 255 (151–501) | 198 (107–255) | 254 (185–426) |
| Number of households | 631 (419–1189) | 294 (229–1202) | 224 (126–381) | 393 (229–1053) |
| Total road length (km) | 8.3 (6.9–10.7) | 5.6 (3.2–9.9) | 4.6 (3.1–6.9) | 6.9 (3.8–9.7) |

<sup>a</sup> Medians with interquartile ranges reported.

Overall, estimated concentrations were lower in 2017-18 compared to 2010 (see **Figures 3-4**), which may be related to a general improvement in air quality for the Mantova province (the larger administrative area including Viadana) during the last decades (ARPA Lombardia 2018). However, it could also be linked to the different weeks during the solar year selected for the two campaigns, which might have captured different seasonal and meteorological conditions.

### 3.6. Comparison between air pollution exposure models

Correlations between estimates of residential exposure attributed to children’s addresses are reported in **Table 4**. Pearson’s r coefficients between pollutants varied widely, from 0.22 (NO_2_ Viadana II vs PM_2.5_ ELAPSE) to 0.86 (NO_2_ vs formaldehyde, Viadana III). Most of the highest correlations were observed between pollutants estimated within the same project, rather than when comparing the same pollutant between different models. To give an example for the models based on routine data, r coefficient for the correlation between NO_2_ and PM_2.5_ from ELAPSE was 0.79, whereas it was 0.42 for PM_2.5_ obtained from ELAPSE and EPISAT.

**Table 4.** Pearson’s r coefficients for the correlation between exposures estimated at children’s residential addresses (n=1,814).

|  |  | Viadana II<br>(2010) |  | ELAPSE<br>(2010) |  |  | EPISAT |  | Viadana III<br>(2017-18) |  |
| --- | --- | --- | --- | --- | --- | --- | --- | --- | --- | --- |
|  |  | NO<sub>2</sub> | Form-<br>aldehyde | NO<sub>2</sub> | PM<sub>2.5</sub> | BC | PM<sub>2.5</sub><br>(2013) | PM<sub>10</sub><br>(2012) | NO<sub>2</sub> | Form-<br>aldehyde |
| Viadana II<br>(2010) | NO<sub>2</sub> | 1 |  |  |  |  |  |  |  |  |
|  | Formaldehyde | 0.42 | 1 |  |  |  |  |  |  |  |
| ELAPSE<br>(2010) | NO<sub>2</sub> | 0.43 | 0.51 | 1 |  |  |  |  |  |  |
|  | PM<sub>2.5</sub> | 0.22 | 0.49 | 0.79 | 1 |  |  |  |  |  |
|  | BC | 0.26 | 0.54 | 0.74 | 0.80 | 1 |  |  |  |  |
| EPISAT | PM<sub>2.5</sub> (2013) | 0.40 | 0.39 | 0.44 | 0.42 | 0.41 | 1 |  |  |  |
|  | PM<sub>10</sub> (2012) | 0.35 | 0.48 | 0.43 | 0.42 | 0.46 | 0.60 | 1 |  |  |
| Viadana III<br>(2017-18) | NO<sub>2</sub> | 0.49 | 0.68 | 0.68 | 0.69 | 0.74 | 0.46 | 0.50 | 1 |  |
|  | Formaldehyde | 0.57 | 0.68 | 0.61 | 0.57 | 0.65 | 0.38 | 0.50 | 0.86 | 1 |

<sup>a</sup> all p<0.001

The three models for NO_2_ consistently detected a maximum concentration at a distance of 1–1.7 km to the industry, followed by a decline as distance increased (**Table 5**), possibly reflecting the pattern of dispersion of nitrogen oxides from the 70-m high chimney. The latter finding is less likely to reflect the local distribution of urban emission sources, since the densities of roads, buildings and population were higher in the buffer closest to the industry. Absolute differences in median NO_2_ exposures between the children at 1–1.7 km and >3.5 km ranged from 2.4 µg/m^3^ (Viadana II) to 5.9 µg/m^3^ (Viadana III). For the other pollutants, estimated exposures decreased with distance: differences in medians between children living at <1 km and ≥3.5 km to the industry were 2.2–2.4 for PM_2.5_ and 5.0 µg/m^3^ for PM_10_. Differences between medians for formaldehyde and BC were small.

**Table 5.** Distribution of exposures estimated at children’s residential addresses, by groups based on quartiles of distance to the industry.

|  | Distance to the industrial plant |  |  |  |  |
| --- | --- | --- | --- | --- | --- |
|  | <1 km<br>(n=454) | 1–1.7 km<br>(n=455) | 1.8–3.4 km<br>(n=452) | 3.5–15.7 km<br>(n=453) | Overall<br>(n=1,814) |
| **Viadana II study (2010)** |  |  |  |  |  |
| NO<sub>2</sub>, µg/m<sup>3</sup> | 17.9 (16.8–18.4) | 19.3 (17.8–20.0) | 18.1 (17.4–18.6) | 16.9 (14.9–17.4) | 17.8 (16.8–18.7) |
| Formaldehyde, µg/m<sup>3</sup> | 2.7 (2.6–2.9) | 2.7 (2.6–2.7) | 2.5 (2.5–2.6) | 2.3 (2.2–2.5) | 2.6 (2.5–2.7) |
| **ELAPSE study (2010)** |  |  |  |  |  |
| NO<sub>2</sub>, µg/m<sup>3</sup> | 28.7 (27.4–29.9) | 29.0 (27.9–29.8) | 28.4 (27.0–29.5) | 24.6 (23.1–26.7) | 28.1 (26.2–29.5) |
| PM<sub>2.5</sub>, µg/m<sup>3</sup> | 26.9 (26.4–27.0) | 26.8 (26.6–26.9) | 26.6 (26.2–26.9) | 24.5 (22.9–26.4) | 26.6 (26.0–26.9) |
| BC, 10<sup>-5</sup> m<sup>-1</sup> | 2.0 (2.0–2.0) | 2.0 (2.0–2.0) | 2.0 (1.9–2.0) | 1.9 (1.8–1.9) | 2.0 (1.9–2.0) |
| **EPISAT study (2012-13)** |  |  |  |  |  |
| PM<sub>2.5</sub> (2013), µg/m<sup>3</sup> | 25.8 (25.0–26.6) | 25.5 (25.2–26.0) | 25.9 (25.1–26.8) | 23.6 (22.5–24.8) | 25.4 (24.6–26.3) |
| PM<sub>10</sub> (2012), µg/m<sup>3</sup> | 38.3 (36.7–40.5) | 37.2 (36.1–39.0) | 36.6 (34.7–38.8) | 33.3 (31.8–34.6) | 36.5 (34.0–38.6) |
| **Viadana III study (2017-18)** |  |  |  |  |  |
| NO<sub>2</sub>, µg/m<sup>3</sup> | 16.5 (15.4–16.8) | 17.1 (16.7–17.2) | 16.0 (13.9–16.8) | 11.2 (10.8–12.1) | 16.2 (13.6–16.9) |
| Formaldehyde, µg/m<sup>3</sup> | 1.7 (1.6–1.8) | 1.8 (1.7–1.8) | 1.7 (1.6–1.8) | 1.6 (1.5–1.6) | 1.7 (1.6–1.8) |

<sup>a</sup> Medians with interquartile range reported. p-values for overall comparisons of concentrations across groups were all <0.001 (Kruskal-Wallis rank test)

In summary, despite the differences in methodology (kriging vs LUR), input data (routine vs passive sampling data), extension (municipality vs national/European), and period (2010-13 vs 2017-18), all the exposure models consistently suggest that children living closer to the chipboard industry (<1.7 km) were more exposed than the children living farther away (>3.5). However, it is important to highlight that, given the proximity between the chipboard industry and the most urbanised areas in Viadana, it is not straightforward to disentangle between emissions from the industry and from other anthropic sources.

### 3.7. Limitations

In agreement with European Directives, we used a continuous analyser based on chemiluminescence as the reference method for NO_2_ measurement (Beelen et al. 2013; Gerboles et al. 2006). Nonetheless we acknowledge that the sample size for validation of passive samplers was small, and missing data in the time-series of air quality data was an additional source of uncertainty. A direct comparison between passive sampling data from the campaigns conducted in 2010 and 2017-18 is hindered by the different number and location of samplers in the monitoring area. Another drawback is that we had no direct PM measurements in the study area. PM are one of the most important emissions from the industry in Viadana, and they convey most of formaldehyde mass that is released during production. They are the most critical component of air pollution in the Po Plain due to their regional distribution and long-term atmospheric persistence (Larsen et al. 2012). LUR models based on routine monitors from country-wide or larger areas perform well in the Po Plain (de Hoogh et al. 2018; Stafoggia et al. 2017) but they may not be ideal to capture variability in industrial pollution, due to the relatively small number of industrial monitoring sites, and the inability of LUR predictors (e.g. land use) to accurately reflect industrial density and specific industrial sources.

## 4. CONCLUSIONS

Using data from passive sampling campaigns conducted in 2017-18, we found that residential exposure to NO_2_ and formaldehyde was higher in proximity to the chipboard industry in Viadana, the most densely urbanised zone in the south of the district. These hot-spots of higher pollutant concentrations persisted from 2010, despite a general air quality improvement in the region. This suggests that spatial gradients in pollutant exposure were stable during the follow-up period of the Viadana III cohort study (2013-2017). Exposure estimates derived from available large-scale LUR models of NO_2_, PM_2.5_ and PM_10_ from routine stations showed associations with distance comparable to our interpolated surfaces. This highlights that people living close to the industry are exposed to a mixture of air pollutants. An implication for Viadana III is that all the previously mentioned models covering the district area (Viadana II, ELAPSE and EPISAT) can be considered equally suitable to derive exposure estimates for the epidemiological analyses. In fact, none of these is a “gold standard” method, and we cannot postulate *a priori* which of the modelled pollutants is the main contributor to specific health effects. Consistency of associations obtained using different air pollution metrics will be key to causal interpretation. Finally, we point that Radiello® tubes may underestimate NO_2_ concentrations, which is important to keep in mind when interpreting association estimates from epidemiological research.

## Data Availability

Air pollution data from passive sampling campaigns conducted in 2017-18 and routine air quality data on NO2 can be found on the ARPA Lombardia website (https://www.arpalombardia.it/). Geocoded addresses of children are not available due to data protection issues. Air pollution data from the Viadana study are available from the corresponding author on reasonable request. Air pollution data from the ELAPSE and EPISAT projects can be requested to the respective study leaders.

## Abbreviations

BC: black carbon
BTEX: benzene, toluene, ethylbenzene, o-xylene, mp-xylene
ELAPSE: Effects of Low-Level Air Pollution: A study in Europe
EPISAT: Dati satellitari ed uso del territorio per la stima delle esposizioni a livello nazionale
LoQ: limit of quantification
LUR: land use regression
NO_2_: nitrogen dioxide
PM: particulate matter
PM_10_: particulate matter with an aerodynamic diameter of 10 μm or less
PM_2.5_: particulate matter with an aerodynamic diameter of 2.5 μm or less

