## Supplementary material for "Spatial variability of nitrogen dioxide and formaldehyde and residential exposure of children in the industrial area of Viadana, Northern Italy": S1

Updated July 17, 2020

**Table S1**. Number of weekly measurements below the limit of quantification for the 25 passive samplers in Viadana (2018-19).

| | $NO_2$ | formaldehyde | benzene | toluene | ethylbenzene | o-xylenes | mp-xylenes | acetaldehyde | benzaldehyde |
|---|---|---|---|---|---|---|---|---|---|
| Limit of quantification (μg/m$^3$) | - | 0.4 | 0.3 | 1.8 | 1.6 | 1.7 | 3.0 | 0.4 | 0.3 |
| Cold season | | | | | | | | | |
| Week n. 1 | 0 | 1 | 0 | 0 | 25 | 25 | 25 | 17 | 25 |
| Week n. 2 | 0 | 0 | 0 | 1 | 25 | 25 | 25 | 0 | 25 |
| Week n. 3 | 0[a] | 4 | 0 | 0 | 25 | 25 | 25 | 9 | 24 |
| Week n. 4 | 0 | 0 | 0 | 2 | 25 | 25 | 25 | 4 | 23 |
| Week n. 5 | 0 | 0 | 1 | 1 | 25 | 25 | 25 | 2 | 24 |
| Warm season | | | | | | | | | |
| Week n. 6 | 0 | 2 | 16 | 22 | 25 | 25 | 25 | 24 | 22 |
| Week n. 7 | 0 | 2 | 25 | 25 | 25 | 25 | 25 | 23 | 25 |
| Week n. 8 | 0 | 6 | 24 | 25 | 25 | 25 | 25 | 24 | 25 |
| Week n. 9 | 0 | 4 | 23 | 25 | 25 | 25 | 25 | 25 | 25 |
| Week n. 10 | 0 | 0 | 25 | 25 | 25 | 25 | 25 | 11 | 10 |

[a] for one sampler (ID 24), the concentration of $NO_2$ at week 3 was imputed as the average between weeks 1, 2, 4 and 5

$NO_2$, nitrogen dioxide

**Figure S1.** Map (panel A) and inset (panel B) of the southern area of the municipality of Viadana (Mantova province). [a]

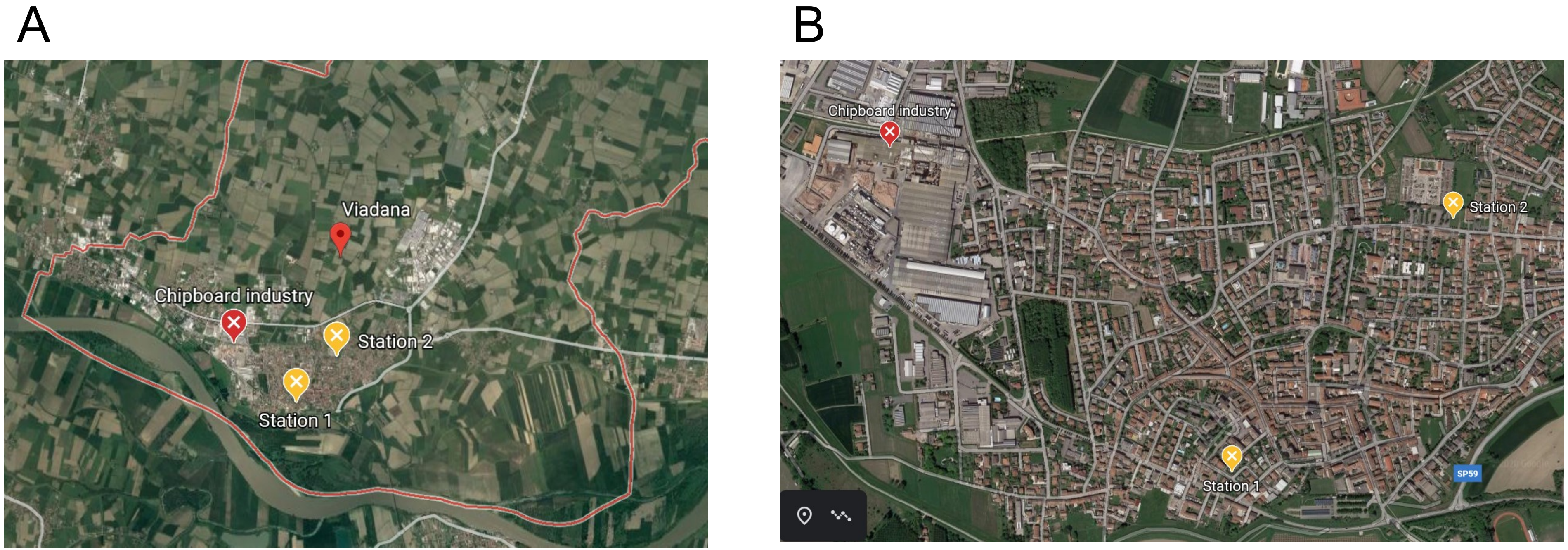


[a] the red line represents the municipality border; red and orange symbols represent the locations of the chipboard industry and the two continuous monitoring stations, respectively (station 1, via Cavalli; station 2, via Airoldi). Maps were obtained using Google Earth™

### Appendix S1. Geocoding procedure

We geocoded residential addresses at baseline in two stages. We first used the World Geocoding Service locator integrated into ArcGIS PRO software (ESRI, Redlands, California). Geocoding was successful if all these conditions were met: matching accuracy score was >90 (range: 0–100, https://pro.arcgis.com/en/pro-app/help/data/geocoding/what-is-included-in-the-geocoded-results-.htm); input and output municipality matched; input street number was recognised, i.e. not removed from the output. Poorly located addresses were geocoded again via outsourcing by EGON Solutions, via Enrico Fermi 13/C, Verona. If also this second stage of geocoding failed, the addresses were excluded from the analysis.

We assessed the agreement between the two geocoding methods (ArcGIS Pro vs EGON), using a stratified random sample of 200 residential addresses (100 addresses from each of the cohorts mentioned in the manuscript). Both methods provided an output in the WGS 1984 coordinate system along with several geocoding quality indicators. This is not a validation analysis since none of the two technique can be considered a "gold standard" *a priori*. As an indicator of agreement we calculated the distance between geocodes obtained for the same address using the two methods. We reported the distribution of pairwise distances and quality indicators for the following samples of addresses:

- Original sample (n=200)
- Selection 1 (n=166): excluding 34 addresses with an ArcGIS match score <90
- Selection 2 (n=160): further excluding six addresses placed at street centreline, i.e. the input street number was not recognised and thus removed from the ArcGIS output. (There were no changes in the output *vs* input municipality, which was a further quality criterion applied for the main analysis. Addresses excluded from selection 2 correspond to the set that was re-geocoded through EGON).
- Selection 3 (n=156): further excluding four addresses that were poorly geocoded also through EGON.

Results from this analysis showed a good agreement between the two geocoding methods (**Table S2**), with a median pairwise distance of 3–4 m whatever the selection considered, and a 75$^{th}$ percentile decreasing from 31.8 m (original sample) to 9.7 m (final selection).

Most of the quality improvement was seen after removing geocodes with a match score <90 (selection 1). Selections at increasing ArcGIS accuracy also showed improved geocoding accuracy according to EGON. In fact, the proportion of matched addresses changed from 74% (original sample) to 79% (final selection).

**Table S2**. Distribution of pairwise distances between geocodes obtained using ArcGIS and EGON, and geocoding quality indicators.

| | Original sample (n=200) | Selection 1 (n=166) | Selection 2 (n=160) | Selection 3 (n=156) |
|---|---|---|---|---|
| Pairwise distance (m) | | | | |
| median | 4.3 | 3.5 | 3.5 | 3.4 |
| 75$^{th}$ percentile | 31.8 | 13.9 | 11.2 | 9.7 |
| 95$^{th}$ percentile | 1836.3 | 372.2 | 298.5 | 98.1 |
| ArcGIS Match score, n (%) | | | | |
| <90 | 34 (17) | 0 (0) | 0 (0) | 0 (0) |
| 90–94 | 17 (8) | 17 (10) | 16 (10) | 15 (10) |
| 95–100 | 149 (75) | 149 (90) | 144 (90) | 141 (90) |
| ArcGIS geocoding accuracy, n (%) | | | | |
| Matched address | 135 (67) | 125 (75) | 125 (78) | 122 (78) |
| Interpolated address | 19 (10) | 18 (11) | 18 (11) | 17 (11) |
| Extrapolated address [a] | 20 (10) | 17 (10) | 17 (11) | 17 (11) |
| Street centreline | 12 (6) | 6 (4) | 0 (0) | 0 (0) |
| Locality centroid/ Point of interest | 14 (7) | 0 (0) | 0 (0) | 0 (0) |
| EGON geocoding accuracy, n (%) | | | | |
| Matched (Navteq) | 147 (74) | 126 (76) | 123 (77) | 123 (79) |
| Interpolated (Navteq) | 33 (16) | 28 (17) | 27 (17) | 27 (17) |
| Interpolated (TeleAtlas) | 12 (6) | 8 (5) | 6 (4) | 6 (4) |
| Municipality / sub-municipality centroid | 8 (4) | 4 (2) | 4 (2) | 0 (0) |
| EGON error report, n (%) | 2 (1) | 2 (1) | 2 (1) | 0 (0) |

[a] input house number exceeds the house number range for the matched street segment

**Figure S2**. Scatterplot of weekly $NO_2$ concentrations measured by the temporary station and a co-located passive sampler ($\mu g/m^3$).[a]

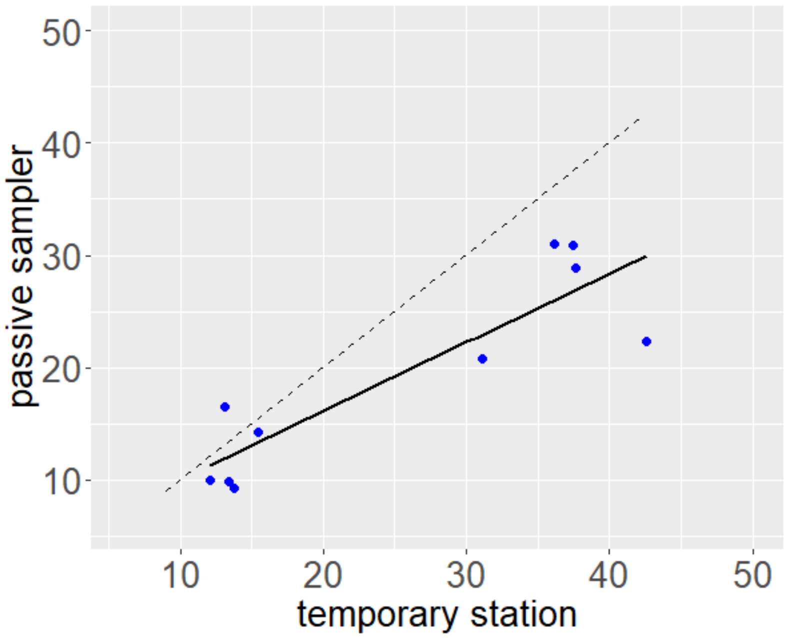


[a] the dashed line is the identity line; the solid line was obtained by linear interpolation:

$NO_2$ passive sampler = 3.92 + 0.61 × $NO_2$ temporary station ($R^2$ = 0.79)

**Table S3**. Distribution of weekly concentrations of benzene, toluene, and acetaldehyde measured by 25 passive samplers in Viadana during the cold season (2017-18).[a]

| Week n. | Starting date (DD/MM/YYYY) | Benzene, $\mu g/m^3$ (mean, SD) | Toluene, $\mu g/m^3$ (mean, SD) | Acetaldehyde, $\mu g/m^3$ (mean, SD) |
|---|---|---|---|---|
| 1 | 21/11/2017 | 1.6 (0.2) | 3.5 (0.5) | most below LoQ |
| 2 | 28/11/2017 | 0.6 (0.1) | 2.4 (0.4) | 1.2 (0.2) |
| 3 | 5/12/2018 | 1.7 (0.2) | 3.0 (0.4) | 0.6 (0.5) |
| 4 | 12/12/2018 | 1.6 (0.2) | 2.5 (0.6) | 0.7 (0.3) |
| 5 | 18/12/2018 | 2.1 (0.5) | 2.8 (0.5) | 0.6 (0.3) |
| Overall (cold season) | | 1.5 (0.1) | 2.8 (0.3) | 0.7 (0.1) |

LoQ, limit of quantification

[a] statistics for the warm season are not reported because more than 50% of the measurements were below the LoQ
